# Deviation from Personalized Blood Pressure Targets Correlates with Worse Outcome after Successful Recanalization

**DOI:** 10.1101/2023.08.10.23293961

**Authors:** Zhe Zhang, Yuehua Pu, Lei Yu, Haiwei Bai, Wanying Duan, Xin Liu, Jingyi Liu, Lina Zheng, Xiao Hu, Xinyi Leng, Yuesong Pan, Nils H. Petersen, Liping Liu

## Abstract

**Background:** Personalized blood pressure (BP) management for acute ischemic stroke patients after successful recanalization lacks evidence. Our study aimed to investigate whether the deviation of BP from cerebral autoregulation (CA) limits, either in duration or burden, is associated with worse outcomes.

**Methods:** We prospectively enrolled patients with acute large anterior circulation artery occlusive stroke who had achieved successful recanalization (mTICI 2b-3). CA was determined by measuring mean velocity index (Mx) using transcranial Doppler sonography (TCD). We calculated the percent time and the burden (defined as the time-BP area) with BP outside the CA limits of each subject within 48 hours after recanalization. The outcomes included unfavorable functional outcome (mRS scored 3-6) at 90 days, early neurological deterioration (END), infarct volume growth at 7 days, and symptomatic intracranial hemorrhage (sICH) within 48 hours.

**Results:** A total of 91 patients were enrolled. Entire failure of autoregulation was found in 31.9% (29 / 91) of patients. The burden with BP outside the CA limits was associated with poor functional outcome at 90 days (a*OR* per 100 h·mmHg 1.29 [95% CI, 1.05 – 1.59] *P*=0.018). Both the percent time and the burden with BP out of the CA limits was correlated with END (a*OR* per 10% time 1.38 [95% CI, 1.05 – 1.83] *P*=0.022, a*OR* per 100h·mmHg 1.22 [95% CI, 1.01 – 1.48] *P*=0.039). The burden of BP that decreased below the CA lower limit was associated with infarct volume growth (adjusted *β* per 100 h·mmHg 5.47 [95% CI, 0.13 – 10.80] *P*=0.045). The percent time that BP exceeded the CA upper limit was associated with sICH (a*OR* per 10% time 1.45 [95% CI, 1.01 – 2.07] *P*=0.043).

**Conclusions:** Both the percent time and burden of BP that deviates from the autoregulation-preserved range correlate with worse outcomes after successful recanalization. Further randomized control trials are warranted to examine the effectiveness of autoregulation-guided BP targets.

---

Endovascular thrombectomy (EVT) improves the prognosis of patients with ischemic stroke due to large-vessel intracranial occlusion with broadened time window[1–4]. However, after successful recanalization, the optimal blood pressure (BP) target is still unknown as several randomized trials (BP-TARGET, ENCHANTED2/MT, BEST-Ⅱ) obtained neutral or even harmful results [5, 6]. Given the high variability in infarct size, location, and risk factors of stroke, a neurophysiological monitoring-based individualized BP management strategy might be an alternative option. Cerebral autoregulation (CA) refers to the capability of cerebral arteries to maintain cerebral blood flow constant within a range of systemic BP changes. It is reasonable to determine an acceptable BP range using CA evaluation in a variety of clinical settings such as traumatic brain injury and intraoperative monitoring [7]. Using near infrared spectroscopy (NIRS), Petersen *et al*. found that the percent time that BP exceeded the upper limits of CA, instead of a fixed target, was associated with unfavorable outcomes in patients who had undergone EVT [8]. Nevertheless, it is unclear whether the degree, in addition to the *duration*, of BP that deviates from CA limits (either above the upper limit or below the lower limit of autoregulation) after successful recanalization, affects outcomes.

In this study, we aimed to test the hypothesis that the *burden* of deviated BP within the first 48 hours after EVT was associated with unfavorable outcomes. The *burden* of deviated BP is defined as the area enclosed by the actual mean arterial pressure (MAP) curve and the upper and/or lower CA-impairment BP thresholds, to reflect the magnitude of BP deviation. Moreover, instead of NIRS that detects only a partial region around middle cerebral artery (MCA) - anterior cerebral artery (ACA) watershed, we used transcranial Doppler sonography (TCD) to assess the autoregulation of MCA which accounts for two-thirds of the cerebral blood supply.

## Materials and Methods

### Study design and subjects

This was a single-center, prospective, observational study. We screened patients who had undergone EVT with acute large artery occlusive stroke from a prospective cohort (*Registry for Critical Care of Acute Stroke in China*, registered in www.chictr.org.cn, identifier ChiCTR1900022154). Patients with large vascular occlusive (LVO) stroke in anterior circulation who had undergone EVT and achieved successful recanalization (modified treatment in cerebral infarction [mTICI] score 2b – 3), aging 18 – 85 years, and premorbid mRS score 0 – 2, were eligible to enroll. We excluded subjects with known any type of hemorrhagic transformation before CA monitoring, isolated ACA territory infarction, infarcts within vertebrobasilar artery territory, aorta dissection, severe internal medical comorbidities (including congestive heart failure, malignant arrhythmia, hepatic failure, and malignant tumors), enrolled in other clinical trials, subclavian artery stenosis, poor acoustic temporal window or cooperation to perform the CA monitoring. The management decisions, including the BP target, followed the current 2018 AHA/ASA and China Stroke Association guidelines [9, 10]. The Ethics Committee of Beijing Tiantan Hospital approved the study (KY2020-140-01). Written informed consent was obtained from the legally authorized representatives of all the subjects.

### Procedures

Upper arm cuff BP measurement as routine care was performed at least every 2 hours for the first 48 hours after EVT. MAP (= [systolic BP – diastolic BP] / 3 + diastolic BP) of each individual was recorded. Within 48 hours after EVT, TCD-based CA monitoring was performed to identify the upper and lower limits of BP with autoregulatory function preserved (**Fig. 1A**). TIME_above-range_ and TIME_below-range_ were the percentage of cumulative time that the MAP respectively exceeded the upper limits and went below the lower limits in the first 48 hours. BURDEN_above-range_ and BURDEN_below-range_ were the area enclosed by the MAP curve above the upper limits, that below the lower CA limits and the corresponding time, respectively. TIME_out-of-range_ and BURDEN_out-of-range_ were the sum of the cumulative time and the area where MAP was out of the range, respectively (**Fig. 1B**). For patients with vasoparalysis (definition see below), BURDEN_out-of-range_ was the area between the MAP curve and the optimal MAP (MAP_OPT_) in 48 hours (**Fig. S1B**).

**Fig. 1.**
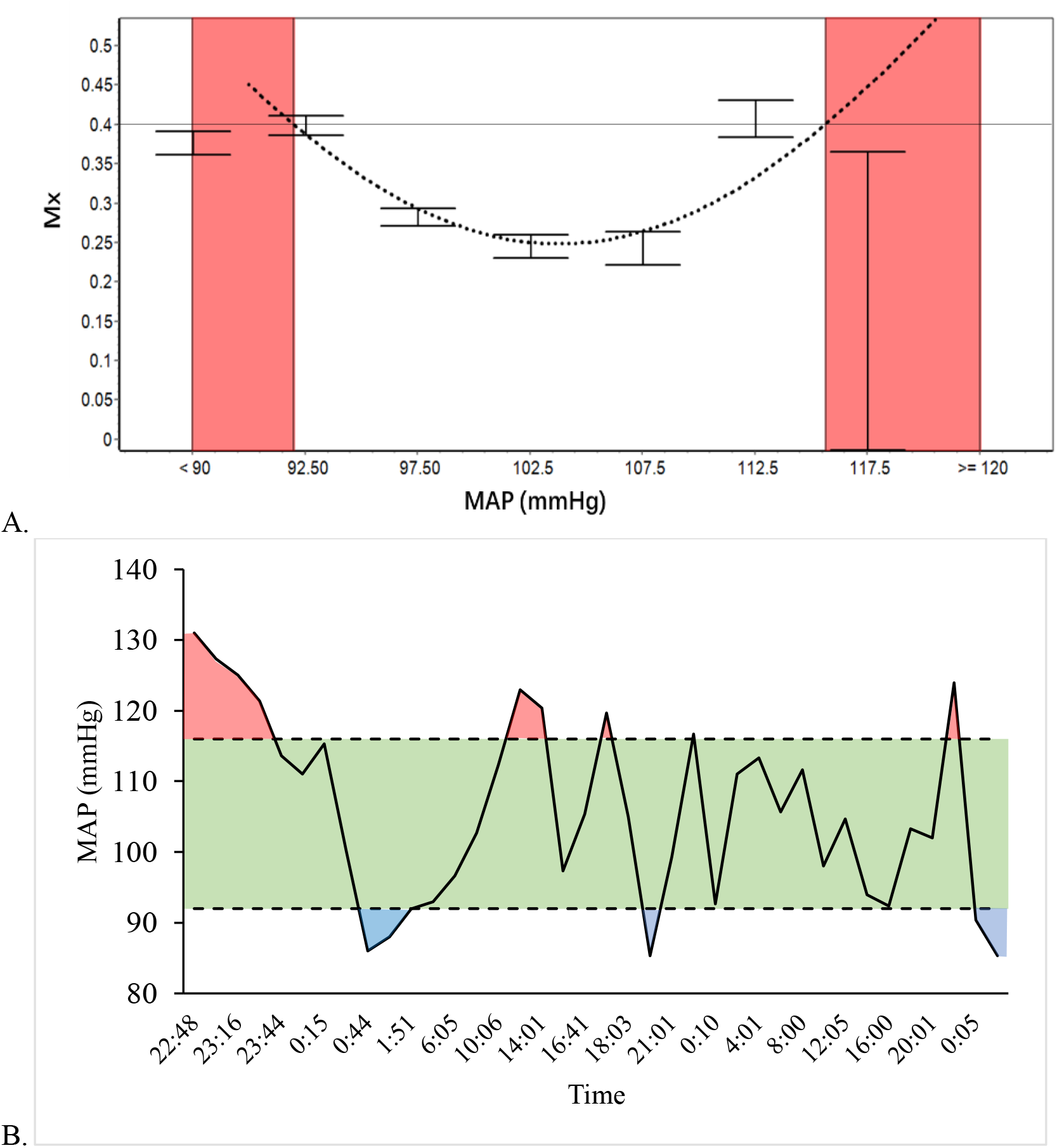
(**A**) A U-shaped Mx-MAP curve could be drawn based on CA monitoring for a patient who had undergone EVT. After applying the Mx=+0.4 threshold, the two intersections of the curve and the threshold line (92 and 116 mmHg) were the upper and the lower limits of autoregulation, respectively. (**B**) MAP fluctuation of the same patient. The green area represented the MAP range in which CA was preserved. The red and blue area were BURDEN_above-range_ and BURDEN_below-range_, respectively. CA indicates cerebral autoregulation; EVT, endovascular therapy; MAP, mean arterial pressure; Mx, mean flow index.

Demographic and clinical data, including the National Institute Stroke Scale (NIHSS) score at admission to the hospital and the 7 days (or at discharge from the neurocritical care unit if less than 7 days), mRS scores at 90 ± 7 days were collected by investigators independent of the CA monitoring. We created ΔNIHSS (=NIHSS scores at 7 days – NIHSS scores at admission) to represent early neurological function changes.

All patients underwent CT angiography and perfusion imaging, or MRI scanning, at baseline and 24 to 48 hours after EVT procedure. A plain CT was performed at 7 days and at any time needed. Estimated by RAPID software (iSchemaView, California of USA), infarct core volume at baseline was the area with an apparent diffusion coefficient threshold of less than 620 × 10^-6^ mm^2^/s on a diffusion-weighted MRI map, or a region with reduced cerebral blood flow less than 30% of that in normal tissue on a CT perfusion scan. A single rater, blind to clinical and CA data, reviewed CT performed 7 days after EVT and manually determined the infarct size. Infarct growth was the difference between the infarct volume at 7 days and the baseline. Based on the ECASS scale [11], hemorrhagic transformation was stratified into three groups: no hemorrhagic transformation (no HT), hemorrhagic infarction (HI), and parenchymal hematoma (PH). The definition of systemic intracranial hemorrhage (sICH) followed the Heidelberg Bleeding Classification [12].

### TCD-based CA monitoring

Subjects underwent TCD monitoring (DWL, Compumedics DWL, Germany) of the bilateral middle cerebral artery (MCA) once or twice within the first 48 hours after EVT. After patients were lying in bed quietly, we placed two 2.5-MHz transducers fitted on a headband over the temporal window. The depth of insonation varied between 50 and 65mm to identify MCA blood flow. MCA flow velocity and arterial BP signals were recorded synchronously for 2 to 4.5 hours. CA was measured by mean velocity index (Mx) using ICM+ software (Cambridge University, UK) [13].

In computing Mx, the two signals were first filtered by a ten-second nonoverlapping average filter. Second, the Mx was computed as a continuous, moving Pearson’s correlation coefficient between thirty consecutive time-averaged MAP values and corresponding MCA blood flow velocity. In this way, each Mx was monitored in a 300-second window. Mx approaches 1 if MCA blood flow passively follows the change of MAP, suggesting CA dysfunction. In contrast, Mx approaches 0 or is negative when CA capacity is preserved. We applied Mx = +0.4 as a cut-point for impaired CA [14]. MAP values were divided into groups of 5 mmHg and plotted against corresponding Mx indices. Then a characteristic U-shaped curve could be drawn. MAP declined below the LLA or exceeded the ULA when Mx increased to the threshold ≥ 0.4 (**Fig. 1A**). The nadir of the U-shaped curve was the MAP_OPT_.

In this study, *vasoparalysis* was a situation where the U-shaped curve entirely exceeded the threshold of Mx = +0.4, indicating that vessels in MCA territory completely lost autoregulatory capacity (**Fig. S1A**).

### Outcomes and hypothesis

The primary outcome was an unfavorable functional independence level (modified Rankin Scale [mRS] scored 3 – 6) at 90 days. The secondary outcomes were early neurological deterioration (END, the NIHSS score at 7 days increased at least 2 scores compared to baseline) [15], ΔNIHSS, infarct volume growth at 7 days compared to baseline, sICH within 48 hours after recanalization, and mRS score at 90 days.

Our hypothesis were as follows: a) the longer TIME_out-of-range_ and the larger BURDEN_out-of-range_ are associated with an increased risk of END, greater disability at 90 day (mRS score 3 – 6), and a higher ΔNIHSS value, b) the longer TIME_above-range_ and the larger BURDEN_above-range_ are associated with a higher risk of sICH within 48 hours, c) the longer TIME_below-range_ and the larger BURDEN_below-range_ are associated with greater infarct volume growth measured at 7 days.

To investigate whether vasoparalysis affects the outcomes, we perform a subgroup analysis by dividing the subjects into vasoparalysis and non-vasoparalysis groups and examining the hypothesis above.

### Statistical methods

Statistical analyses were performed using StataMP 17.0 (Stata Corporation, Texas). Data were reported as mean ± standard deviation (*SD*) if they obeyed normal distribution or median (interquartile range [*IQR*]) if not. We used the Wilcoxon rank-sum test, *χ*^2^ test, *t*-test, Fisher’s exact test for comparing two unadjusted samples, Kruskal-Wallis test for more than two independent samples. We applied the Spearman correlation coefficient to examine the correlation between the two groups. Odds ratios (*OR*s) of dichotomous variables, and correlation coefficients (*β*) for continuous variables were estimated using a logistic and linear regression model, respectively. *OR*s and *β* values were adjusted for age, admission NIHSS score, time from symptom onset to reperfusion, admission MAP and the Alberta Stroke Program Early CT score (ASPECTS). *P* values < 0.05 were considered statistically significant.

## Results

### Subject characteristics

Between May 5, 2020, and Feb 21, 2022, a total of 200 patients were screened, and 120 TCD-based CA monitoring sessions were performed among the 91 enrolled subjects (**Fig. S2**). The CA monitoring began at 21.7 (*IQR* 14.1 – 35.4) hours after recanalization, with 3.2 ± 1.1 hours of artifact-free recording. The average number of BP measurements was 36.6 ± 8.2, with average MAP 97 ± 10 mmHg. The median of TIME_out-of-range_ and BURDEN_out-of-range_ was 75.9% (*IQR* 40.4% - 99.1%) and 295.0 (*IQR* 133.5 - 488.1) h • mmHg, respectively. All the subjects successfully completed follow-up. More detailed characteristics are summarized in **Table 1**.

**Table 1.** Patient characteristics.

|  |  |
| --- | --- |
| Total patients included | 91 |
| Age, yr, mean ± SD | 59.8 ± 11.5 |
| Male, n (%) | 76 (83.5) |
| Clinical, laboratory, and imaging data at admission to the hospital |  |
| Admission NIHSS, median (*IQR*) | 13 (11 – 17) |
| Admission SBP mmHg, mean ± SD | 148 ± 26 |
| Admission DBP mmHg, mean ± SD | 86 ± 18 |
| Admission MAP, mmHg, mean ± SD | 107 ± 20 |
| Admission glucose, mmol/L, mean ± SD | 7.5 ± 2.6 |
| Left-sided infarct, n (%) | 52 (57.1) |
| ASPECTS score, median (*IQR*) | 8 (7 – 9) |
| Occlusion on angiography, n (%) |  |
| ICA (C1 – 3) | 29 (31.9) |
| ICA (C4 – 7) | 13 (14.3) |
| MCA (M1 – M2) | 49 (53.8) |
| Medical history, n (%) |  |
| Hypertension | 60 (65.9) |
| Coronary artery disease | 20 (22.0) |
| Atrial fibrillation | 25 (27.5) |
| Dislipidemia | 23 (25.3) |
| Diabetes mellitus | 26 (28.6) |
| Smoking | 56 (61.5) |
| TIA or stroke | 32 (35.2) |
| Stroke subtype, n (%) |  |
| Large artery atherosclerotic | 63 (69.2) |
| Cardioembolism | 22 (24.2) |
| Others† | 6 (6.6) |
| Recanalization treatment |  |
| Intravenous tPA, n (%) | 33 (36.3) |
| Stent retrieval attempts, median (*IQR*) | 1 (0 – 2) |
| Time from symptom onset to reperfusion, min, median (*IQR*) | 509 (350 – 780) |
| TICI 2b, n (%) | 19 (20.9) |
| TICI 3, n (%) | 72 (79.1) |
| Post-procedural blood pressure |  |
| Number of measurements, mean ± SD | 36.6 ± 8.2 |
| SBP, mmHg, mean ± SD | 135 ± 14 |
| DBP, mmHg, mean ± SD | 77 ± 10 |
| MAP, mmHg, mean ± SD | 97 ± 10 |
| Coefficient of variation of SBP, mean ± SD‡ | 10.8 ± 2.7 |
| Coefficient of variation of DBP, mean ± SD‡ | 13.4 ± 3.9 |
| Coefficient of variation of MAP, mean ± SD‡ | 10.9 ± 3.1 |
| TCD-based CA monitoring |  |
| Time from reperfusion to CA monitoring initiation, min, median (*IQR*) | 1304 (847 – 2127) |
| Dynamic parameters |  |
| TIME<sub>out-of-range</sub>, %, median (*IQR*) | 75.9 (40.4 – 99.1) |
| BURDEN<sub>out-of-range</sub>, mmHg·hr, median (*IQR*) | 295.0 (133.5 – 488.1) |
| TIME<sub>above-range</sub>, %, median (*IQR*) | 13.5 (0 – 42.9) |
| BURDEN<sub>above-range</sub>, mmHg·hr, median (*IQR*) | 25.1 (0 – 182.7) |
| TIME<sub>below-range</sub>, %, median (*IQR*) | 37.4 (8.7 – 68.5) |
| BURDEN<sub>below-range</sub>, mmHg·hr, median (*IQR*) | 111.1 (13.8 – 305.4) |
| Hemorrhagic transformation, n (%) | 35 (38.5) |
| HI 1 | 8 (8.8) |
| HI 2 | 18 (19.8) |
| PH 1 | 3 (3.3) |
| PH 2 | 6 (6.6) |
| sICH | 6 (6.6) |
| END, n (%) | 16 (17.6) |
| Infarct growth at 7 days, mL, median (*IQR*) | 10 (2 – 60) |
| 90-d mRS 3-6, n (%) | 56 (61.5) |

ASPECTS indicates Alberta Stroke Program Early CT Score; DBP, diastolic blood pressure; END, early neurological deterioration; HI, hemorrhagic infarction; ICA, internal carotid artery; IQR, interquartile range; MAP, mean arterial pressure; MCA, middle cerebral artery; mRS, modified Rankin Scale; TICI, Thrombolysis in Cerebral Infarction; NIHSS, National Institute of Health Stroke Scale; PH, parenchymal hematoma; SBP, systolic blood pressure; SD, standard deviation; sICH, symptomatic intracerebral hemorrhage; TIA, transient ischemic attack; tPA, tissue plasminogen activator.
† Including one subject with internal carotid artery dissection, and five cryptogenic.
‡ Coefficient of variation ( $=SD_{BP} / \text{Mean}_{BP} \times 100$ ) represents the variability of blood pressure [32].

### Outcomes

The median of ΔNIHSS was -3 (*IQR* -8 – 0). ΔNIHSS was associated with TIME_out-of-range_ and BURDEN_out-of-range_ (*P*=0.010, 0.016, respectively) (**Fig. 2 A, B**). Linear regression model demonstrated a correlation between BURDEN_out-of-range_ and ΔNIHSS (adjusted *β* per 100h • mmHg 0.51 [95% CI, 0.07 – 0.96] *P*=0.024) (**Table 3**). Sixteen (17.6%) subjects had END. Patients with END had longer TIME_out-of-range_ (96.8% [*IQR* 84.5% – 100%] versus 67.3% [*IQR* 34.1% – 97.3%] *P*=0.005) and larger BURDEN_out-of-range_ (447.6 [*IQR* 313.0 – 511.3] h • mmHg *versus* 247.9 [*IQR* 88.3 – 470.4] h • mmHg *P*=0.012). In multivariate Logistic regression model analysis, both TIME_out-of-range_ and BURDEN_out-of-range_ were noted as independent risk factors of END (a*OR* per 10% time 1.38 [95% CI, 1.05 – 1.83] *P*=0.022, a*OR* per 100h • mmHg 1.22 [95% CI, 1.01 – 1.48] *P*=0.039) (**Table 2**).

**Fig. 2.**
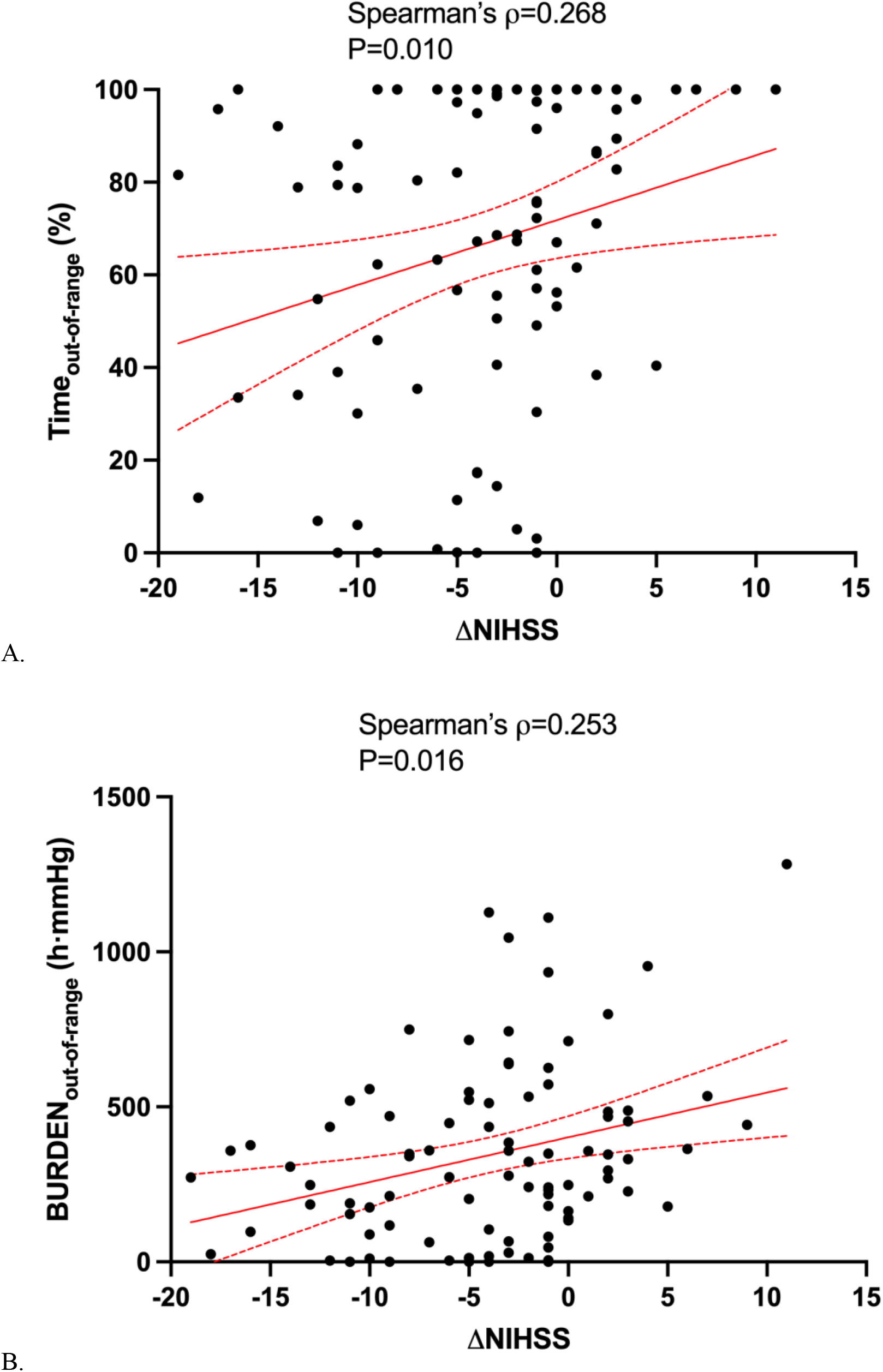
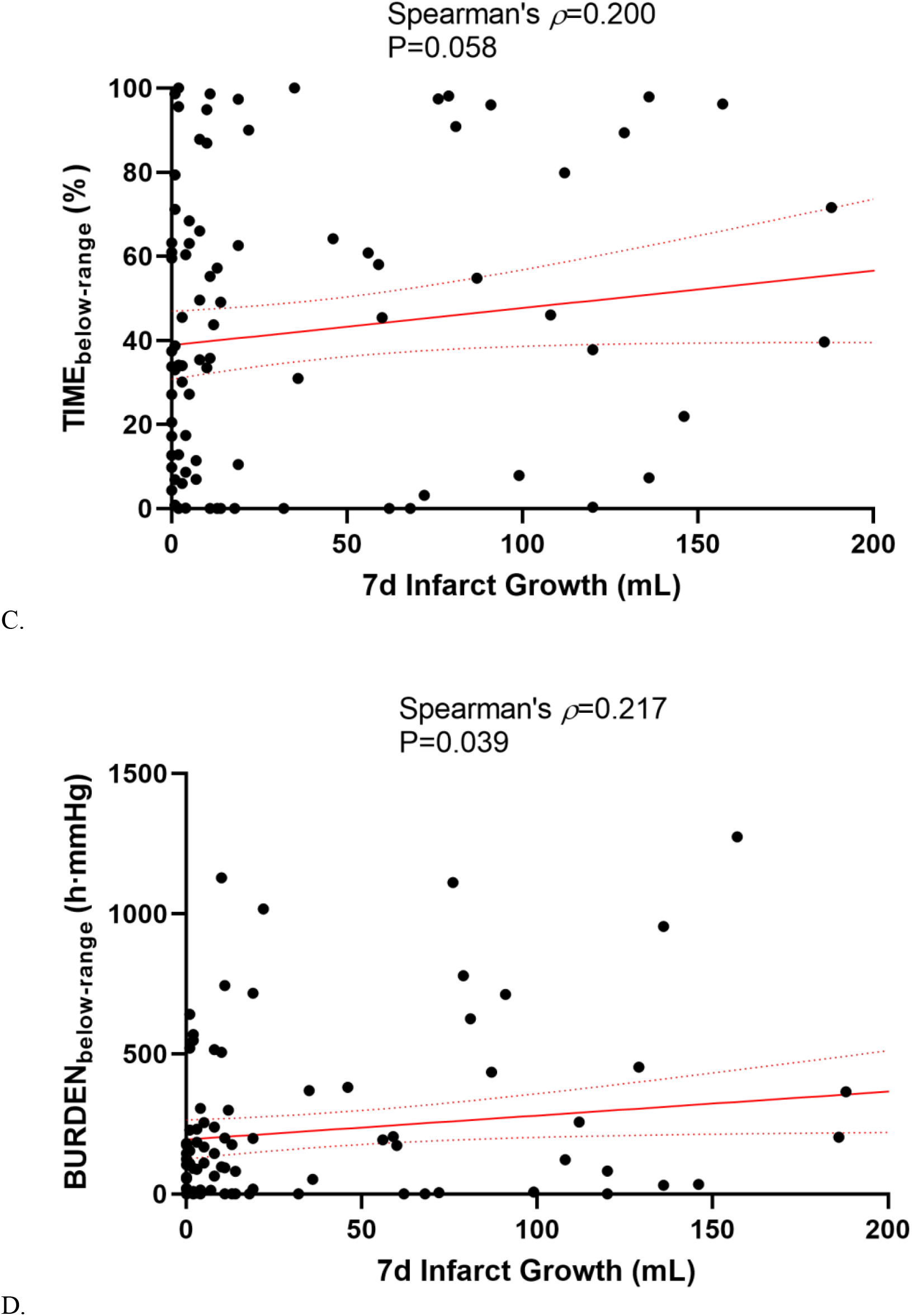
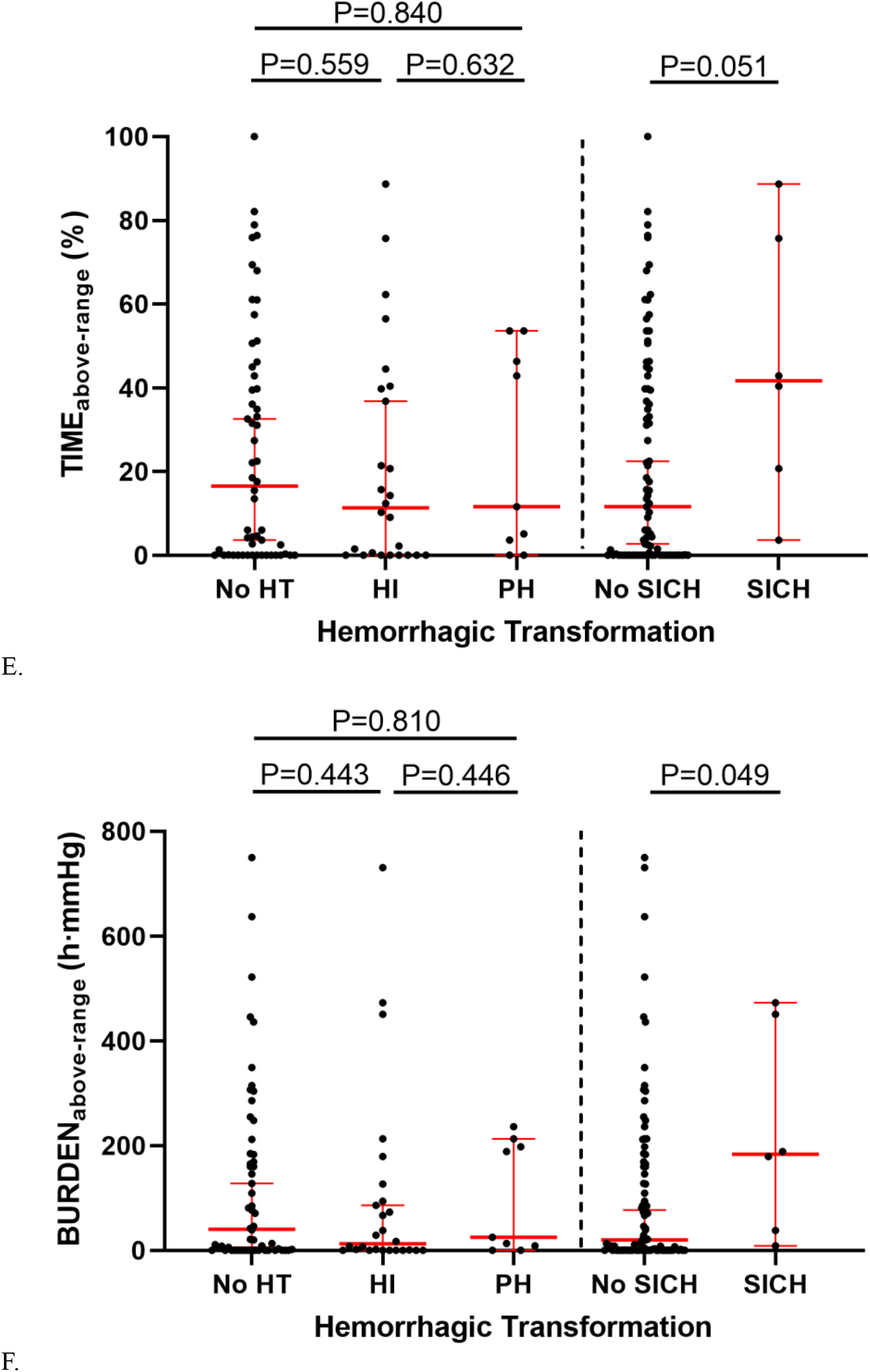
(**A, B**) Both the TIME_out-of-range_ and the BURDEN_out-of-range_ were associated with ΔNIHSS (=NIHSS scores at 7 days – NIHSS at admission). (**C, D**) The BURDEN_below-range_ was associated with infarct growth within 7 days, while the correlation between TIME^below-range^ and infarct growth was statistically insignificant. **(E, F)** Patients with sICH had a higher TIME_above-range_ and BURDEN_above-range_ but the differences were of marginal significance. HI indicates hemorrhagic infarction; HT, hemorrhagic transformation; NIHSS, National Institute of Health Stroke Scale; PH, parenchymal hematoma; sICH, symptomatic intracranial hemorrhage.

**Table 2.** Deviation of blood pressure from autoregulation limits with outcomes using Logistic regression model.

| Variables | *OR* | 95% CI | *P* value | *aOR*† | 95% CI | *P* value |
| --- | --- | --- | --- | --- | --- | --- |
| END |  |  |  |  |  |  |
| Every 10% time increase in TIME<sub>out-of-range</sub> | 1.38 | 1.06 – 1.79 | 0.015* | 1.38 | 1.05 – 1.83 | 0.022* |
| Every 100h·mmHg increase in BURDEN<sub>out-of-range</sub> | 1.22 | 1.02 – 1.46 | 0.027* | 1.22 | 1.01 – 1.48 | 0.039* |
| sICH |  |  |  |  |  |  |
| Every 10% increase in TIME<sub>above-range</sub> | 1.31 | 0.99 – 1.74 | 0.059 | 1.45 | 1.01 – 2.07 | 0.043* |
| Every 100h·mmHg increase in BURDEN<sub>above-range</sub> | 1.33 | 0.92 – 1.92 | 0.127 | 1.57 | 0.98 – 2.53 | 0.061 |
| 90-day functional outcome (mRS 3 – 6) |  |  |  |  |  |  |
| Every 10% increase in TIME<sub>out-of-range</sub> | 1.17 | 1.03 – 1.33 | 0.016* | 1.14 | 0.99 – 1.31 | 0.067 |
| Every 100h·mmHg increase in BURDEN<sub>out-of-range</sub> | 1.37 | 1.11 – 1.68 | 0.003* | 1.29 | 1.05 – 1.59 | 0.018* |

†Adjusted for age, admission NIHSS score, time from symptom onset to reperfusion, admission MAP, and ASPECTS score.
\* $P < 0.05$ .
ASPECTS indicates Alberta Stroke Program Early CT Score; CI, confidential interval; END, early neurological deterioration; MAP, mean arterial pressure; mRS, modified Rankin Scale; NIHSS, National Institute of Health Stroke Scale; *OR*, odds ratio; sICH, symptomatic intracranial hemorrhage.

**Table 3.**
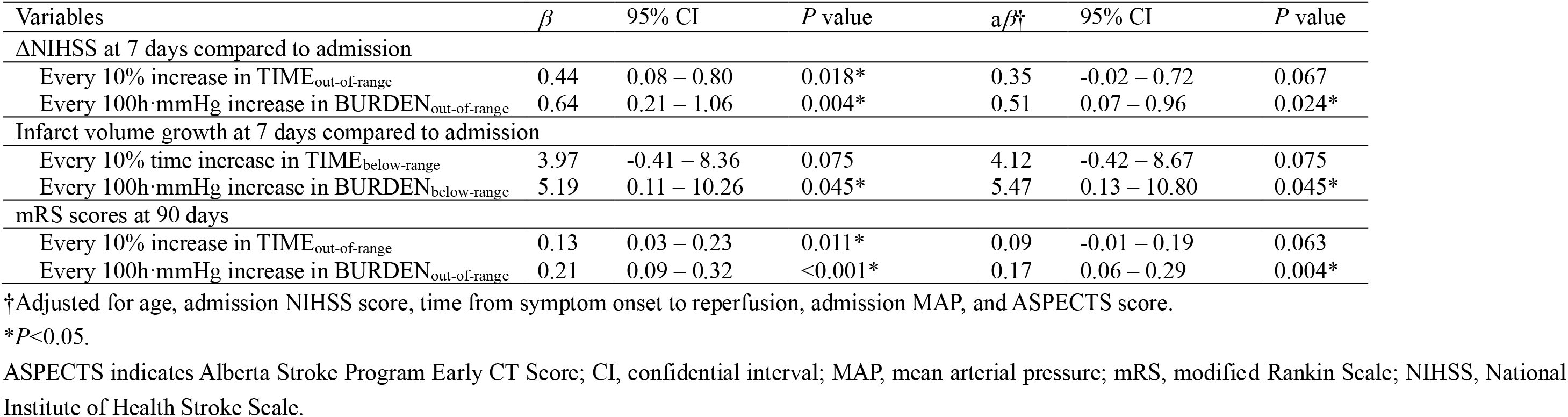
Deviation of blood pressure from autoregulation limits with outcomes using a linear regression model.

The median volume of infarct growth at 7 days was 20 (*IQR* 2-60) mL. The BURDEN_below-range_ was associated with infarct volume growth (*P*=0.039). The correlation between TIME_below-range_ and infarct progress at 7 days was trended in a similar direction but was not statistically significant (*P*=0.058) (**Fig. 2 C, D**). Linear regression model demonstrated that only the BURDEN_below-range_ was associated with infarct growth (a*β* per 100 h·mmHg 5.47 [95% CI, 0.13 – 10.80] *P*=0.045), whereas the correlation between TIME_below-range_ and infarct progress was not statistically significant (**Table 3**).

Thirty-five (38.5%) subjects had hemorrhagic transformation with various types (**Table 1**), and six of them were symptomatic. Patients with sICH had a higher TIME_above-range_ and BURDEN_above-range_ but with marginal significance (41.7% [*IQR* 20.7% – 75.7%] *versus* 11.6% [*IQR* 0 – 39.8%] *P*=0.051, 183.8 [*IQR* 38.3 – 450.9]h • mmHg *versus* 19.7 [*IQR* 0 – 165.0] h • mmHg *P*=0.049, respectively) (**Fig. 2 E, F**). In the adjusted analysis, only TIME_above-range_ was associated with sICH (a*OR* per 10% time 1.45 [95% CI, 1.01 – 2.07] *P*=0.043) (**Table 2**).

Fifty-six (61.5%) subjects developed unfavorable functional outcome (mRS scored 3-6) at 90 days. Both TIME_out-of-range_ and BURDEN_out-of-range_ were higher in those with poor outcome (84.5% [*IQR* 56.5% – 100%] *versus* 62.3% [*IQR* 17.4% – 86.7%] *P*=0.012, 358.0 [*IQR* 193.9 –541.1] h • mmHg *versus* 212.8 [*IQR* 29.7 – 358.5] h • mmHg *P*=0.003, respectively). In multivariate analysis, BURDEN_out-of-range_ was associated with mRS score (a*β* per 100 h • mmHg 0.17 [95% CI 0.06 – 0.29] *P*=0.004), as well as poor functional outcome (a*OR* per 100 h • mmHg 1.29 [95% CI, 1.05 – 1.59] *P*=0.018) (**Fig. 3 A, B**, **Table 2** and **3**).

**Fig. 3.**
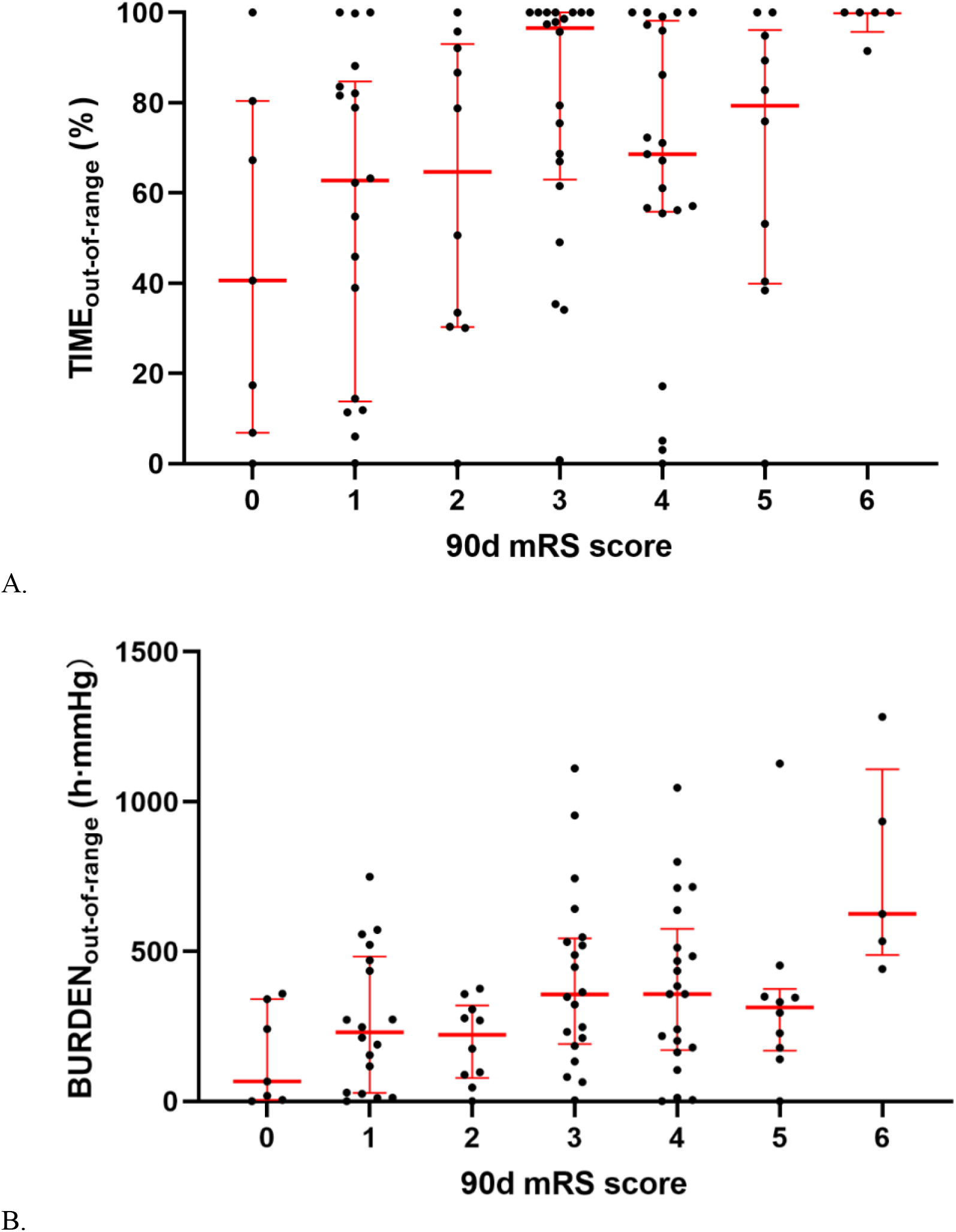
(**A, B**) Both the TIME_out-of-range_ and the BURDEN_out-of-range_ demonstrated an increasing trend with higher 90-day mRS score. mRS indicates modified Rankin Scale.

### Vasoparalysis

Vasoparalysis was found in 31.9% (29 out of 91) subjects and accounted for 28.3% (34 out of 120) of the total TCD-based CA monitoring. Patients with vasoparalysis had higher ASPECTS scores than those without (median 9 [*IQR* 8 – 9] versus 7 [*IQR* 6 – 8], *P*<0.001). The differences in admission NIHSS scores and duration between reperfusion to monitoring were insignificant (**Supplementary Table 1**).

On the subgroup analysis, none of the TIME or BURDEN had statistically significant effect or correlation with the outcomes, although a marginal effect was found in BURDEN_out-of-range_ among non-vasoparalysis subjects on END (a*OR* per 100 h • mmHg 1.45 [95% CI 0.99 – 2.11] *P*=0.054), and a marginal association was noticed in BURDEN_out-of-range_ among those who had vasoparalysis with 90-day mRS score (a*β* per 100h·mmHg 0.20 [95% CI -0.01 – 0.42] *P*=0.059) (**Supplementary Table 2** and **3**).

## Discussion

This study demonstrated that deviation of BP from a personalized, autoregulation-based target (either exceeding or decreasing below) after LVO stroke with successful recanalization was associated with an increased risk of infarct progression and hemorrhagic transformation, which further affected acute and long-term functional outcomes. This BP target can be identified by TCD-based CA monitoring after the thrombectomy procedure. If blood pressure exceeds the upper limit or falls below the lower limit, it can cause hyper- or hypoperfusion. As TCD is widely used in inpatient care for stroke, our study suggests the potential benefits of personalized BP management with generalizability.

According to the randomized trials on mechanical revascularization, nearly half of patients who had undergone EVT had unfavorable outcomes [1]. Observational studies denoted that both high and low BP values at the acute stage were associated with increased early and late mortality [16]. To date, there are three randomized controlled trials (BP-TARGET, ENCHANTED2/MT, and BEST-Ⅱ) to assess whether BP lowering treatment post successful recanalization could improve the prognosis. Intensive control of systolic BP to fixed targets <130mmHg or < 120mmHg failed to increase the odds toward favorable outcome (mRS score 0-2) at 90 days [5], or even demonstrated harm [6]. As previously stated, having a single, fixed target for blood pressure may not be appropriate for all patients due to differences in their history of hypertension and collateral circulation, which affects their cerebral perfusion needs. In addition to stratified ranges, other characteristics of BP, such as variability and trajectory, have attracted attention. Higher BP variability after successful recanalization was associated with worse long-term outcomes, especially in those with poor collateral compensation [17, 18]. Patients with high, moderate-to-high, and high-to-moderate systolic BP trajectories were more likely to have unfavorable outcomes [19]. We found that every 100 h·mmHg increase in the magnitude (described as the *burden* in this research) that BP deviated from CA limits had a 1.29-fold increase in the odds of a poor functional outcome at 90 days. Our result provides a possible explanation for the findings above. The changes or fluctuation in BP may lead to an increase in the burden of BP outside the CA-preserved range, which further causes poor long-term outcomes.

We found that both the time and the burden with BP out of CA range were associated with END. Further analysis suggested that this association might be owing to infarct volume growth and sICH, which were possibly caused by increased BURDEN_below-range_ and TIME_above-range_, respectively. The burden below the lower CA limit was noticed to be related to infarct volume growth, suggesting that compared to the time, the magnitude of the BP reduction contributed more to worsening the cerebral perfusion less than 17 – 18ml/(100g • min), resulting in permanent ischemia [20]. As systemic BP drops below the lower limits of CA, deterioration in cerebral hypoperfusion would reduce microemboli clearance, and lead to infarct progression after successful recanalization [21, 22]. Thus, pressor therapy in progressive acute ischemic stroke might be warranted if the BP is below the lower limit of autoregulation.

In contrast, the relationship between sICH and BP above the upper limit was time-dependent. Upadhyaya *et al.* found in post-EVT patients (*n*=103), lower systolic BP (<130mmHg) was not associated with decreased risk of sICH, although as many as 69% of patients had shown evidence of blood-brain barrier (BBB) disruption [23]. Nevertheless, research based on larger cohorts and meta-analysis unlocked the connection between the sICH and higher systolic BP [24–26]. Apart from cerebral hemodynamic factors, mechanisms including post-ischemia inflammation also contribute to the pathogenesis of hemorrhagic transformation by altering BBB permeability [27]. Our findings and the literature suggested that the elevation in hydrostatic pressure driven by systemic BP might not simply follow a linear relationship with the amount or composition of efflux through BBB.

Our study first reported that after successful recanalization, cerebral vessels could be so severely injured that there was no such BP interval in which autoregulation was preserved. In this circumstance, cerebral blood flow is always pressure-passive. We use the old-fashioned but appropriate term *vasoparalysis* to describe this entire failure of autoregulation. We found that patients with vasoparalysis had higher ASPECTS scores before EVT, suggesting that the large infarct size may be responsible [28]. Accumulated acidic metabolites and adenosine in infarct tissue could dilate cerebral arterioles [29, 30], causing cerebral autoregulation failure. Meanwhile, the monitoring sessions which detected vasoparalysis were not closer to recanalization in time than those detected no vasoparalysis. This demonstrates that vasoparalysis may not recover quickly, at least in the short postoperative period (48 hours). In patients with vasoparalysis, BP management turns more difficult because of the increased risk of both hemorrhage and infarct progress. A possible solution to this dilemma is to bring BP as close to MAP_OPT_ as possible to minimize the BURDEN_out-of-range_, since marginal (though statistically insignificant) correlation was revealed between BURDEN_out-of-range_ and 90-day mRS score in vasoparalysis subgroup (**supplementary Table 3**).

A key difference between ours and Petersen *et al.*’s work [8] is the CA evaluation methods. As mentioned above, NIRS might have difficulties in reflecting the whole-brain autoregulation by attaching probes on the forehead (around the MCA-ACA watershed) to measure changes in regional brain tissue oxygenation. Besides its wide accessibility, the advantage of TCD which used in this study is the capability to evaluate the autoregulatory function responsible for up to two-thirds of the cerebral hemisphere. Because ischemic stroke is usually not a general but rather focal brain disease, expansion in the detectable area makes it more sensitive to identify impaired autoregulation. This explains why we discovered vasoparalysis in nearly one-third of the subjects, and the TIME_out-of-range_ was much higher compared to Petersen *et al*.’s measurements (∼35%).

Our study has several limitations. First, TCD, as a measure of CA, requires good temporal window and cooperation from patients. The prospect of extended duration monitoring raises theoretical concerns regarding potential mechanical compromise to cerebral structures and cerebrovasculature [31]. These shortcomings make continuous monitoring impossible. Second, low frequency, “spot” measurements of BP and CA made TIME and BURDEN calculation relatively inaccurate. Third, for the baseline NIHSS score, we used the one at admission rather than post-procedural or at the time of enrollment, making it unable to reflect the improvement from successful EVT. This situation arose because a substantial number of patients had received general anesthesia and/or intubation, resulting in challenges when conducting precise neurological assessments. Fourth, the study featured a moderate sample size, and while it yielded significant results for its primary and secondary outcomes, the subgroup analysis focused on vasoparalysis did not achieve statistical significance.

## Conclusions

Our results show that after successful recanalization, the percent time and burden of BP outside the autoregulation-preserved range respectively correlate with short-term and long-term clinical outcomes. Infarct progress is associated with the magnitude of BP below the lower CA limit, whereas the relationship between sICH and BP above the upper limit is time-dependent. It is suggested that not only percent time, but also burden of BP out of CA-preserved range should be minimized. Further prospective, randomized control trials are warranted to examine the effectiveness of autoregulation-guided BP management strategy.

## Data Availability

All data referenced in this manuscript are available upon reasonable request.

## Acknowledgement

We thank engineers from Beijing Beike Digital Medical Technology Co., Ltd. for technical support and all the clinical and nursing staff in the Neurocritical Care Unit of Beijing Tiantan Hospital for assistance in this study.

